# SARS-CoV-2 seroprevalence and implications for population immunity: Evidence from two Health and Demographic Surveillance System sites in Kenya, February-June 2022

**DOI:** 10.1101/2022.10.10.22280824

**Authors:** EW Kagucia, AK Ziraba, J Nyagwange, B Kutima, M Kimani, D Akech, M Ng’oda, A Sigilai, D Mugo, H Karanja, J Gitonga, A Karani, M Toroitich, B Karia, M Otiende, A Njeri, R Aman, P Amoth, M Mwangangi, K Kasera, W Ng’ang’a, S Voller, LI Ochola-Oyier, C Bottomley, A Nyaguara, PK Munywoki, G Bigogo, E Maitha, S Uyoga, KE Gallagher, AO Etyang, E Barasa, J Mwangangi, P Bejon, IMO Adetifa, GM Warimwe, JAG Scott, A Agweyu

## Abstract

**Background:** Up-to-date SARS-CoV-2 antibody seroprevalence estimates are important for informing public health planning, including priorities for Coronavirus disease 2019 (COVID-19) vaccination programs. We sought to estimate infection- and vaccination-induced SARS-CoV-2 antibody seroprevalence within representative samples of the Kenyan population approximately two years into the COVID-19 pandemic and approximately one year after rollout of the national COVID-19 vaccination program.

**Methods:** We conducted cross-sectional serosurveys within random, age-stratified samples of Kilifi Health and Demographic Surveillance System (HDSS) and Nairobi Urban HDSS residents. Anti-spike (anti-S) immunoglobulin G (IgG) and anti-nucleoprotein (anti-N) IgG were measured using validated in-house ELISAs. Target-specific Bayesian population-weighted seroprevalence was calculated overall, by sex and by age, with adjustment for test performance as appropriate. Anti-S IgG concentrations were estimated with reference to the WHO International Standard (IS) for anti-SARS-CoV-2 immunoglobulin and their reverse cumulative distributions plotted.

**Results:** Between February and June 2022, 852 and 851 individuals within the Kilifi HDSS and the Nairobi Urban HDSS, respectively, were sampled. Only 11.0% (95% confidence interval [CI] 9.0-13.3) of all Kilifi HDSS participants and 33.4% (95%CI 30.2-36.6) of all Nairobi Urban HDSS participants had received any doses of COVID-19 vaccine. Population-weighted anti-S IgG seroprevalence was 69.1% (95% credible interval [CrI] 65.8-72.3) within the Kilifi HDSS and 88.5% (95%CrI 86.1-90.6) within the Nairobi Urban HDSS. Among COVID-unvaccinated residents of the Kilifi HDSS and Nairobi Urban HDSS, it was 66.7% (95%CrI 63.3-70.0) and 85.3% (95%CrI 82.1-88.2), respectively. Population-weighted, test-adjusted anti-N IgG seroprevalence within the Kilifi HDSS was 53.5% (95%CrI 46.5-61.1) and 65.5% (95%CrI 56.0-75.6) within the Nairobi Urban HDSS. The prevalence of anti-N antibodies was similar in vaccinated and unvaccinated subgroups in both HDSS populations. Anti-S IgG concentrations were significantly lower among Kilifi HDSS residents than among Nairobi Urban HDSS residents (p< 0.001).

**Conclusions:** Approximately, 7 in 10 Kilifi residents and 9 in 10 Nairobi residents were seropositive for anti-S IgG by May 2022 and June 2022, respectively. Given COVID-19 vaccination coverage, anti-S IgG seropositivity among COVID-unvaccinated individuals, and anti-N IgG seroprevalence, population-level anti-S IgG seroprevalence was predominantly derived from infection. Interventions to improve COVID-19 vaccination uptake should be targeted to individuals in rural Kenya who are at high risk of severe COVID-19.

## Introduction

Serosurveillance for SARS-CoV-2 antibodies emerged as an important tool for estimating the cumulative incidence of SARS-CoV-2 infection in the early phase of the COVID-19 pandemic. It was particularly important in settings with low COVID-19 testing levels, including many countries in Africa.^1^ In these settings, cumulative incidence revealed by SARS-CoV-2 serosurveillance far outstripped the rate of infection inferred from case detection.^2^ In the current context, SARS-CoV-2 serosurveillance remains important for assessing temporal changes in seroprevalence, including waning, and identifying potential gaps in population immunity to inform priorities for COVID-19 prevention measures, including vaccination.

By June 2020, roughly three months after the first confirmed case of COVID-19 in Kenya, seroprevalence of SARS-CoV-2 anti-spike (anti-S) immunoglobulin G (IgG) was 4% among blood donors.^3^ Continued serosurveillance among blood donors in Kenya showed temporal increases in anti-S IgG seroprevalence, reaching 48% by March 2021,^4^ one year after identification of the first confirmed SARS-CoV-2 infection locally which also coincided with the rollout of the COVID-19 vaccination program by the Government of Kenya. The COVID-19 vaccination program initially targeted frontline workers, older adults aged ≥58 years, and younger adults with comorbidities. It soon opened up to all adults, then to children aged ≥15 years in November 2021, and currently targets individuals aged ≥12 years. To date, the national program has included the following COVID-19 vaccines: Oxford/ AstraZeneca (Covishield), Pfizer (BNT162b2), Moderna (mRNA-1273), Johnson & Johnson (Ad26.CoV2.S) and Sinopharm (BBIBP-CorV). By 31 July 2022, full vaccination coverage among adults nationally was 34% and 11% among children aged 15-17 years, albeit with substantial heterogeneity at the sub-national level.^5^

Randomly selected residents of health and demographic surveillance systems (HDSS) lack the selection biases inherent in blood transfusion donors and provide a more representative sample of the general population. By May 2021, two months after initial rollout of COVID-19 vaccine, serosurveillance among an age-stratified random sample of residents of three HDSS sites found anti-S IgG seroprevalence ranging from 25% to 50%.^6^ We undertook a repeat serosurvey at two of these three HDSS sites to assess infection-and vaccination-induced seroprevalence within the general population approximately two years into the pandemic and one year after rollout of the COVID-19 vaccination program.

## Methods

### Study design and participants

A cross-sectional survey was conducted at each of two sites, the Kilifi HDSS and the Nairobi Urban HDSS. The characteristics of these HDSS sites have been described in detail previously.^7,8^ The Kilifi HDSS is located in a rural area within Kilifi County in south-eastern coastal Kenya, while the Nairobi Urban HDSS is located within Nairobi County, the capital city of Kenya (**Figure S1**). As of the most recent census data available, the population of the Kilifi HDSS was 308,581 (April 2022) and about 90,000 at the Nairobi Urban HDSS (October 2021).

We used similar methods as previous SARS-CoV-2 serosurveys at the sites.^6^ In brief, a random age-stratified sample of 850 participants was drawn from the respective HDSS site population registers. The sample drawn at each site was independent of that drawn for the surveys conducted in 2020-21. The simple random sample included 100 children in each five-year age band <15 years, 50 individuals in each five-year age band between 15 and 64 years, and 50 adults aged ≥65 years. Individuals were eligible to participate in the study if they were resident in the respective HDSS, had no contraindication for blood sample collection, and provided consent. Individuals were considered not contactable after three unsuccessful attempts to visit them. A random replacement sample was drawn when all individuals in the original sample of 850 had been contacted per study operating procedures but fewer than 850 participants enrolled.

The study research protocol was aligned to the World Health Organization (WHO) UNITY methods for COVID-19 serosurveillance studies.^9^ Ethical approval was obtained from the Kenya Medical Research Institute Scientific and Ethics Review Unit (KEMRI/SERU/CGMR-C/203/4085), the Oxford Tropical Research Ethics Committee (44-20), and the London School of Hygiene and Tropical Medicine Research Ethics Committee (26950).

### Participant consent statement

Written parental/guardian consent was obtained for participants aged <18 years, accompanied by written assent for children aged 13-17 years. Written informed consent was obtained from participants aged ≥18 years.

### Data and sample collection

Sociodemographic (e.g., age, sex, location of residence) information and medical history data (e.g., recent COVID-like symptoms, COVID-19 vaccination history, previous confirmed SARS-CoV-2 infection) were collected from each participant (**Supplement**). COVID-19 vaccination status was ascertained using either official records (i.e., COVID-19 vaccination certificate or text message confirmation from the national COVID-19 vaccine registry) or verbal report. A single 2mL (children aged <5 years) or 5mL (individuals aged ≥5 years) venous blood sample was collected from each participant and labelled with a unique identifier.

### Laboratory testing

Plasma was extracted from venous blood samples and tested for anti-S IgG to identify either infection- or vaccination-induced antibody response, and for anti-nucleoprotein (anti-N) IgG, to identify infection-induced antibody response.^10^ Testing was performed using validated KEMRI-Wellcome Trust Research Programme ELISAs. Sensitivity and specificity for the anti-S IgG ELISAs were, respectively, 93% (95% confidence interval [CI] 88-96) and 99% (95%CI 98-99).^3^ Sensitivity and specificity for the anti-N IgG ELISA were 83% (95%CI 76-88) and 91% (86-95). Target-specific IgG positivity was defined as a ratio of the sample optical density (OD) over negative control OD >2. The (WHO International Standard (IS) for anti-SARS-CoV-2 immunoglobulin (NIBSC code 20/136) was included in each anti-S IgG ELISA run and used to calculate sample-specific binding antibody concentrations in binding antibody units per millilitre (BAU/mL). A binding antibody concentration of 1000 BAU/mL was assigned to the WHO IS, as recommended by NIBSC.^11^ Sample-specific binding antibody concentrations were calculated by dividing each sample-specific OD ratio by the run-specific IS OD ratio after which the quotient was multiplied by 1000.

### Statistical analysis

The target sample size of 850 participants was sufficient to measure antibody seroprevalence of 50% with an associated 95% confidence interval (CI) of ±3%.

COVID-19 vaccine coverage was calculated as the proportion of individuals reporting vaccination divided by the entire sample. Exact binomial 95%CIs were also calculated. Coverage with ≥1 doses of COVID-19 vaccine and full vaccination coverage were estimated overall and restricted to participants aged ≥15 years. Full vaccination was defined as receipt of at least one dose of Johnson & Johnson vaccine or receipt of ≥2 doses of other COVID-19 vaccines.

Crude seroprevalence was calculated as the proportion of seropositive samples over all samples, along with exact binomial 95%CIs. Because the population sample did not represent the age-structure of the target population, Bayesian population weighting was performed to generate population-weighted seroprevalence and associated 95% credible intervals (CrI). Anti-S IgG seroprevalence within the entire sample was not adjusted for test performance as validation of the ELISA was performed among unvaccinated individuals; for example, adjustment for test-performance using current sensitivity and specificity estimates would overestimate anti-S seroprevalence if in truth assay sensitivity is higher among COVID-vaccinated individuals. To estimate infection-induced seroprevalence, Bayesian population-weighted anti-S IgG seroprevalence was estimated among the subset of participants who were COVID-unvaccinated. Anti-S IgG seroprevalence among COVID-unvaccinated participants was not adjusted for test-performance for ease of interpretation given that anti-S IgG seroprevalence within the entire sample was similarly not adjusted for test performance, as described above. To assess the magnitude of inherent assay bias, Bayesian test-performance adjusted anti-S seroprevalence was also estimated among only COVID-unvaccinated participants.

To assess the contribution of infection to seroprevalence levels, population-weighted and test-adjusted anti-N IgG seroprevalence was calculated separately for COVID-vaccinated and COVID-unvaccinated participants. Given limited use of inactivated COVID vaccines in Kenya, which can induce anti-N IgG responses,^12^ it was deemed appropriate to adjust the anti-N seroprevalence estimates within the entire sample.

For each set of seroprevalence data, seroprevalence was computed overall, by sex and by seven age strata (<16; 16-24; 25-34; 35-44; 45-54; 55-64; ≥65 years). These age strata were selected to allow comparison to seroprevalence data from the same HDSS sites in 2020-2021.^6^

Reverse cumulative distribution curves (RCDC) for anti-S IgG concentrations were plotted. Kolmogorov-Smirnov tests were used to test the equality of the overall distributions across sites, as well as across sex, age group and COVID-19 vaccination status within each site. A Bonferroni correction was applied to the p-value for the six pairwise age group comparisons using children aged <16 years as the reference group (i.e., p≤ 0.008 was considered significant). The comparison of the distribution of antibodies for COVID-unvaccinated *vs* COVID-vaccinated individuals was restricted to those aged ≥15 years to minimize confounding by age given that children aged <12 years were not eligible for COVID-19 vaccination. The proportion of individuals with anti-S IgG ≥154 BAU/mL was calculated; ≥154 BAU/mL has been proposed as a threshold of protection against infection with wild-type SARS-CoV-2 following COVID-19 vaccination.^13^

Population-weighting and adjustments for test-performance were performed in R with RStan and all other analyses were performed using Stata.

## Results

At the Kilifi HDSS site, a total of 1389 potential participants were visited at the household, of whom 1168 were present. Of those, 853 (73.0%) provided consent to participate in the study, and 852 were sampled between 15 February and 08 May 2022 (median 23 March 2022). A total of 1404 potential participants were visited at the Nairobi Urban HDSS site, of whom 978 were present. Of those present, 852 (87.1%) provided consent and 851 were sampled between 08 March and 22 June 2022 (median 01 May 2022; **Figure 1**). The timing of sample collection at the Kilifi HDSS site occurred just after the Omicron (BA.1) wave, while the sample collection period at the Nairobi Urban HDSS site began after the Omicron BA.1 wave and continued up to the first half of the Omicron BA.4/BA.5 wave (**Figure S2**).

**Figure 1.**
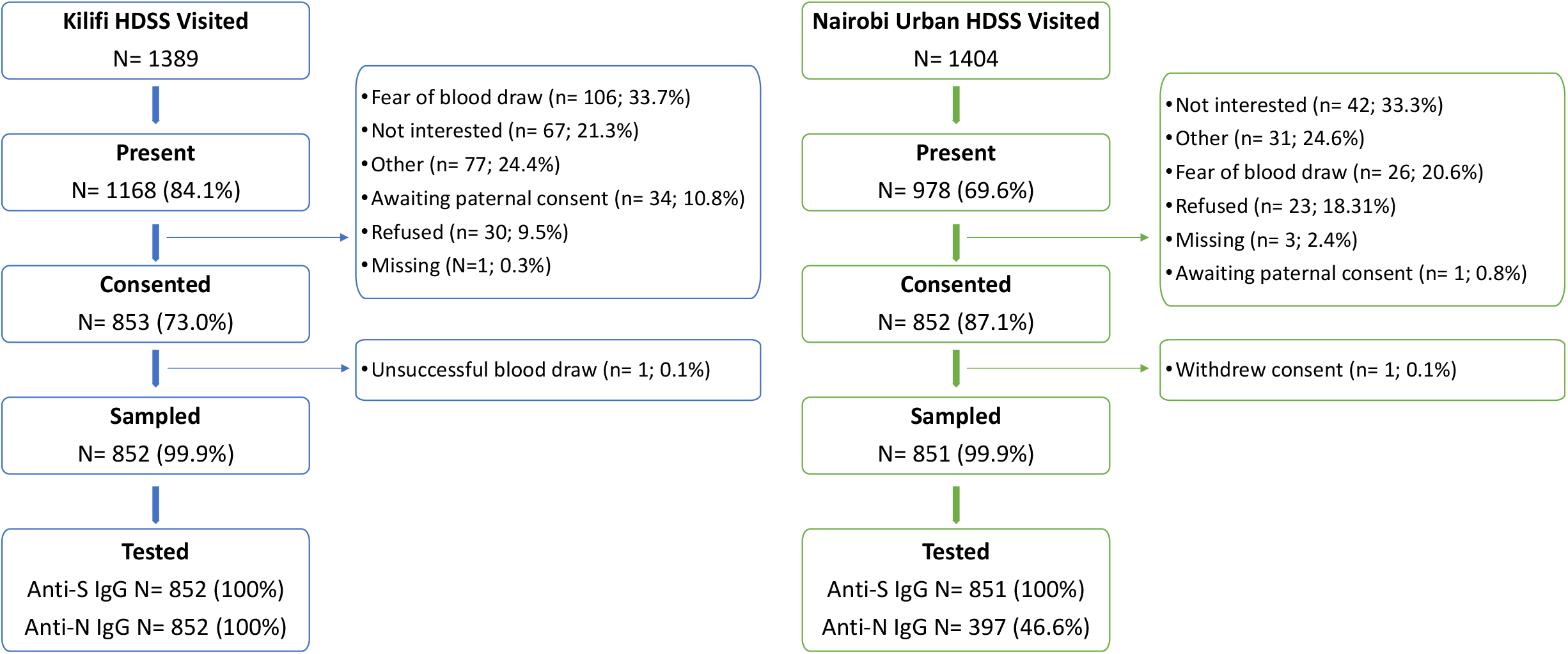
Study participant flow

The median age of participants at both sites was 27 years (interquartile range, 10-49 at Kilifi HDSS and 11-48 at Nairobi Urban HDSS). Among participants reporting COVID-19 vaccine receipt, vaccination was ascertained using an official record in 28.7% (27 of 94) and 27.8% (79 of 284) within the Kilifi HDSS and the Nairobi Urban HDSS, respectively. COVID-19 vaccine uptake was significantly lower among residents of the Kilifi HDSS than among Nairobi Urban HDSS residents. Coverage with ≥1 doses among all enrolled participants was 11.0% (95%CI 9.0-13.3) and 33.4% (95%CI 30.2-36.6) at the Kilifi HDSS site and the Nairobi Urban HDSS site, respectively. When restricted to participants aged ≥15 years of age, coverage within the Kilifi HDSS and Nairobi Urban HDSS, respectively, was: 16.8% (94 of 559; 95%CI 13.8-20.2) and 50.4% (284 of 563; 95%CI 46.2-54.6) for ≥1 doses of COVID-19 vaccine; 9.8% (55 of 559; 95%CI 7.4-12.6) and 37.5% (211 of 563; 95%CI 33.5-41.6) for full vaccination. Among the 852 Kilifi HDSS participants, 177 (20.7%) reported ≥1 COVID-like symptoms within two weeks prior to sample collection, of whom 1.7% (3 of 177) required hospitalization. The proportion reporting ≥1 recent COVID-like symptoms among the 851 Nairobi Urban HDSS participants was 414 (48.6%) of whom 1.4% (6 of 414) were hospitalized (**Table S1**). The top three symptoms at both sites were cough, headache and runny nose and were most commonly reported among children aged <16 years (**Figures S3 and S4**).

### Anti-S IgG seroprevalence and concentrations

Population-weighted anti-S IgG seroprevalence was 69.1% (95%CrI 65.8-72.3) among all 852 participants at the Kilifi HDSS site, which was significantly lower than the 88.5% (95%CrI 86.1-90.6) seroprevalence among all 851 participants at the Nairobi Urban HDSS site. Within the Nairobi Urban HDSS site, anti-S IgG seroprevalence was significantly lower among children aged <16 years (81.4%; 95%CrI 76.0-85.6) than among individuals aged 16-24 years (91.7%; 95%CI 86.2-95.8), 35-44 years (94.6%; 95%CI 90.1-97.8); 45-54 years (94.0%; 95%CI 89.4-97.4) and 55-64 years (95.2%; 95%CI 90.2-98.2). Seroprevalence was significantly lower across all age and sex strata among Kilifi HDSS residents compared to Nairobi Urban HDSS residents (**Table 1**).

**Table 1.**
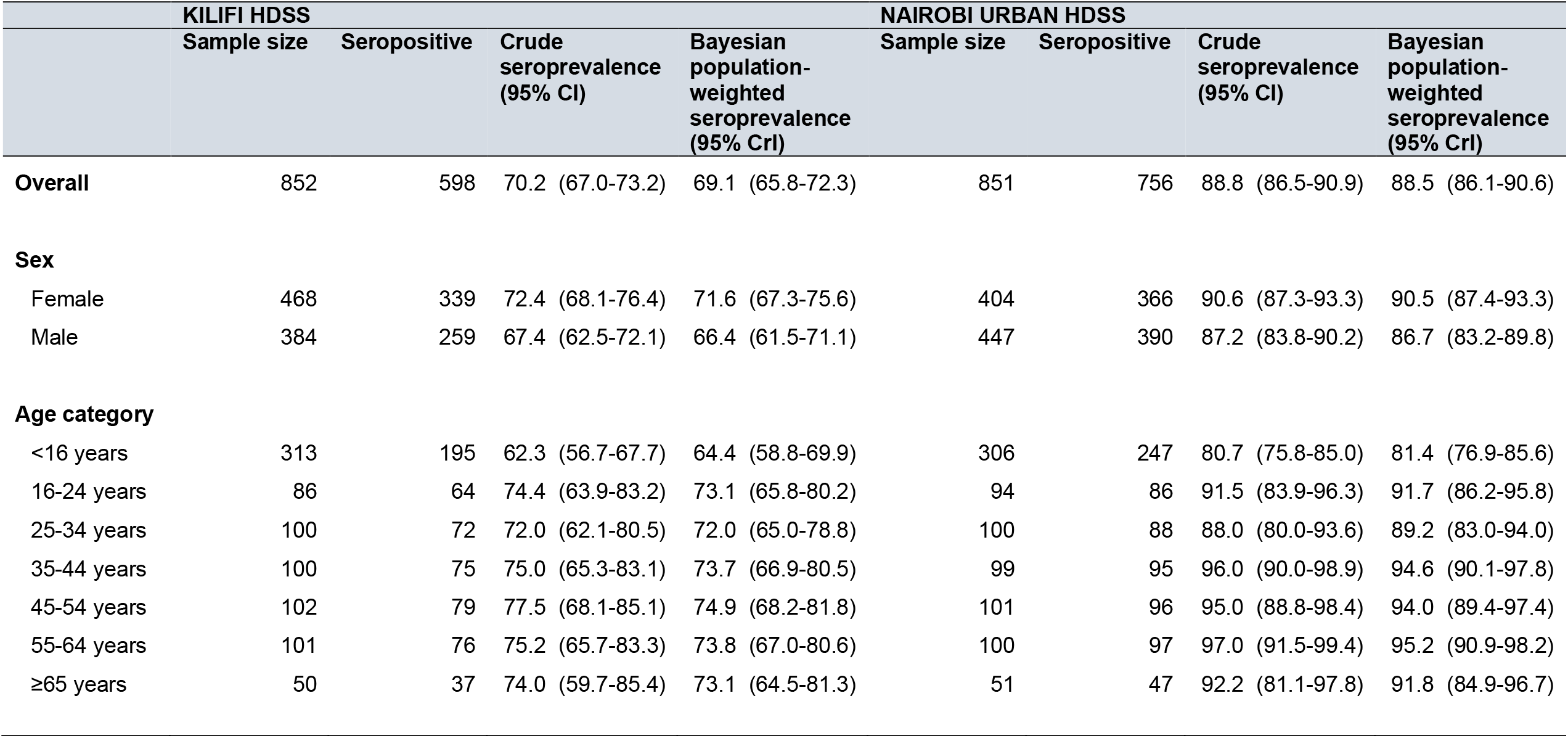
Population-weighted **anti-spike IgG** seroprevalence among **all study participants** by site, sex and age category.

Anti-S IgG concentrations were significantly lower within the Kilifi HDSS than within the Nairobi Urban HDSS (p< 0.001). The median anti-S IgG concentration was ≥328 BAU/mL among Kilifi HDSS residents and ≥799 BAU/ML among Nairobi Urban HDSS residents. The proportion of individuals with anti-spike IgG ≥154 BAU/mL was 62.6% and 85.8% within the Kilifi HDSS and Nairobi Urban HDSS, respectively (**Figure 2 & Table S2**). Within the Kilifi HDSS, antibody concentrations were significantly lower among children <16 years than among all other age groups (p≤ 0.007) other than adults aged ≥65 years. Antibody concentrations for children aged <16 years within the Nairobi Urban HDSS were significantly lower than among all other age groups (p< 0.001). At each of the sites, antibody concentrations were significantly lower among COVID-unvaccinated participants aged ≥15 years than among COVID-vaccinated participants (p< 0.001; **Figure S5; Table S2**).

**Figure 2.**
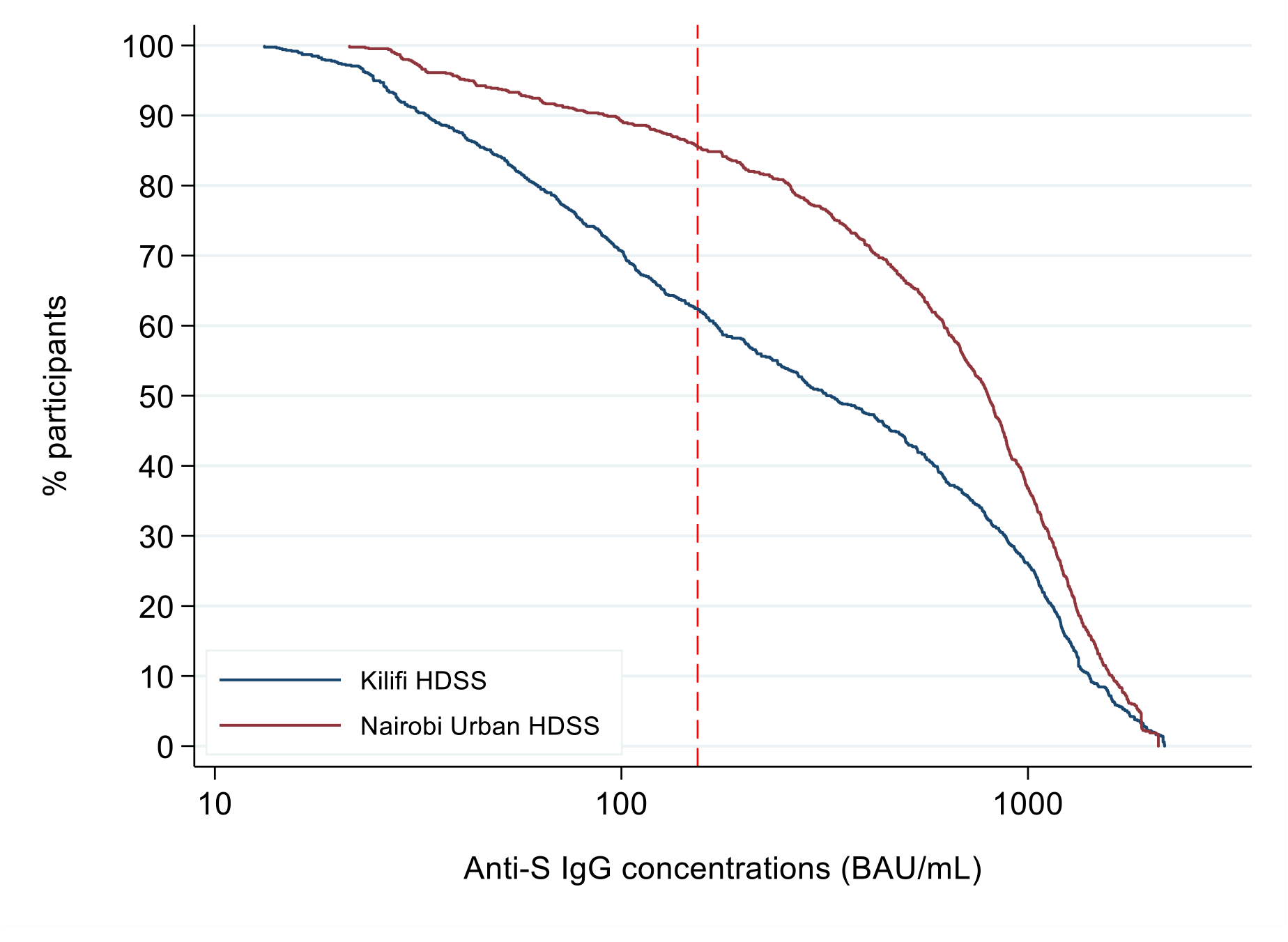
Reverse cumulative distribution curves of anti-S IgG concentrations within the Kilifi HDSS and Nairobi Urban HDSS. The red vertical line represents an antibody concentration of 154 BAU/mL.

COVID-unvaccinated participants represented 89.0% (758 of 852) and 66.6% (567 of 851) of participants at the Kilifi HDSS site and the Nairobi Urban HDSS site, respectively. Population-weighted anti-S IgG seroprevalence among them was 66.7% (95%CrI 63.3-70.0) within the Kilifi HDSS and 85.3% (95%CrI 82.1-88.2) within the Nairobi Urban HDSS. Seroprevalence was significantly lower among Kilifi HDSS residents than Nairobi Urban HDSS residents overall as well as by sex and age (i.e., non-overlapping CrIs; **Table 2**). Seroprevalence estimates among COVID-unvaccinated participants were slightly higher after adjustment for test performance, but not significantly so (**Table S3**).

**Table 2.** Population-weighted and test-adjusted **anti-spike IgG** seroprevalence among **COVID-unvaccinated** individuals by site, sex and age category

| **KILIFI HDSS** | **Sample size** | **Seropositive** | **Crude seroprevalence**<br>**(95% CI)** | **Bayesian population-**<br>**weighted seroprevalence**<br>**(95% CrI)** |
| --- | --- | --- | --- | --- |
| Overall | 758 | 508 | 67.0 (63.5-70.4) | 66.7 (63.3-70.0) |
| **Sex** |  |  |  |  |
| Female | 422 | 293 | 69.4 (64.8-73.8) | 69.3 (64.7-73.7) |
| Male | 336 | 215 | 64.0 (58.6-69.1) | 63.8 (58.6-68.7) |
| **Age category** |  |  |  |  |
| <16 years | 312 | 194 | 62.2 (56.5-67.6) | 64.4 (59.1-69.0) |
| 16-24 years | 74 | 53 | 71.6 (59.9-81.5) | 68.8 (62.5-76.0) |
| 25-34 years | 86 | 59 | 68.6 (57.7-78.2) | 68.1 (61.6-75.0) |
| 35-44 years | 88 | 63 | 71.6 (61.0-80.7) | 69.2 (63.1-76.1) |
| 45-54 years | 81 | 58 | 71.6 (60.5-81.1) | 69.1 (62.8-76.3) |
| 55-64 years | 75 | 51 | 68.0 (56.2-78.3) | 68.0 (61.1-74.6) |
| ≥65 years | 42 | 30 | 71.4 (55.4-84.3) | 68.9 (61.7-77.0) |
| **NAIROBI URBAN**<br>**HDSS** | **Sample size** | **Seropositive** | **Crude seroprevalence**<br>**(95% CI)** | **Bayesian population-**<br>**weighted seroprevalence**<br>**(95% CrI)** |
| Overall | 567 | 479 | 84.5 (81.2-87.4) | 85.3 (82.1-88.2) |
| **Sex** |  |  |  |  |
| Female | 283 | 248 | 87.6 (83.2-91.2) | 88.5 (84.5-92.0) |
| Male | 284 | 231 | 81.3 (76.3-85.7) | 82.5 (77.8-86.7) |
| **Age category** |  |  |  |  |
| <16 years | 300 | 242 | 80.7 (75.7-85.0) | 81.4 (76.8-85.5) |
| 16-24 years | 60 | 52 | 86.7 (75.4-94.1) | 86.8 (79.7-92.7) |
| 25-34 years | 65 | 55 | 84.6 (73.5-92.4) | 85.8 (78.7-91.8) |
| 35-44 years | 48 | 45 | 93.8 (82.8-98.7) | 89.7 (82.5-95.9) |
| 45-54 years | 40 | 35 | 87.5 (73.2-95.8) | 86.3 (77.7-93.3) |
| 55-64 years | 39 | 38 | 97.4 (86.5-99.9) | 90.5 (82.8-97.1) |
| ≥65 years | 15 | 12 | 80.0 (51.9-95.7) | 84.7 (72.0-93.2) |

### Anti-N IgG seroprevalence

The analysis includes anti-N seroprevalence data for all participants at the Kilifi HDSS site (N= 852) and for 47% (397 of 851) participants at the Nairobi Urban HDSS site (**Figure 1**). Population-weighted, test-adjusted anti-N IgG seroprevalence within the Kilifi HDSS was 53.5% (95%CrI 46.5-61.1) and 65.5% (95%CrI 56.0-75.6) within the Nairobi Urban HDSS. At the Kilifi HDSS site, anti-N IgG seroprevalence among COVID-unvaccinated participants was 54.2% (95%CrI 46.9-53.2) and 58.0% (95%CrI 40.1-76.5) among COVID-vaccinated participants. At the Nairobi Urban HDSS site, it was 66.8% (95%CrI 56.1-77.4) among COVID-unvaccinated participants and 78.8% (95%CrI 59.3-99.6) among COVID-vaccinated participants (**Table 3**).

**Table 3.**
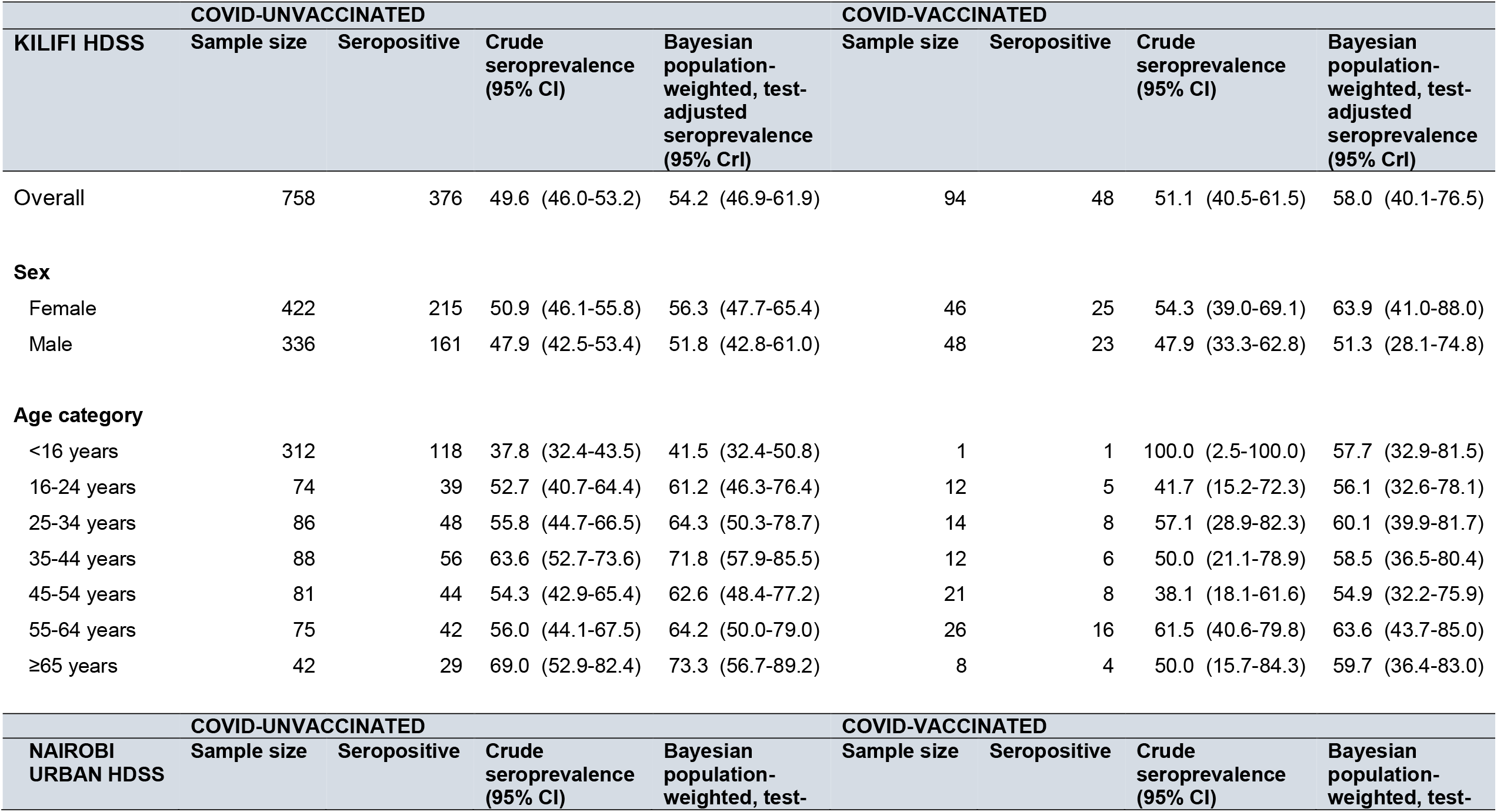
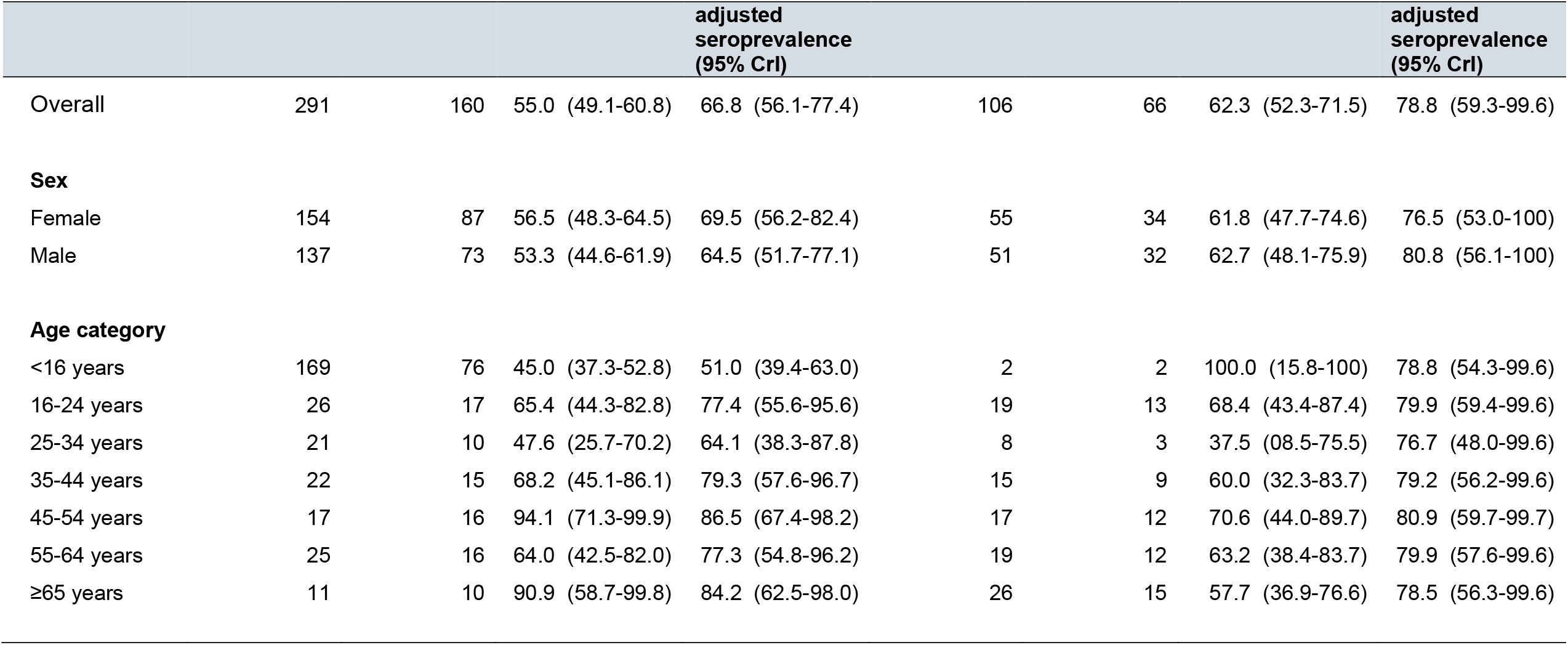
Population-weighted and test-adjusted **anti-nucleoprotein IgG** seroprevalence among **COVID-unvaccinated** and **COVID-vaccinated** study participants by site, sex and age category

## Discussion

We estimate anti-S IgG seroprevalence of 69% by May 2022 in Kilifi and 88% by June 2022 in Nairobi. Yet, only about 1 in 10 of study participants in Kilifi and about 3 in 10 in Nairobi had received any doses of COVID-19 vaccine, indicating the substantial contribution of natural infection to anti-S IgG seroprevalence. Indeed, anti-S IgG seroprevalence among COVID-unvaccinated participants – who made up the majority of the study sample – was 67% and 85% in Kilifi and Nairobi, respectively, indicative of infection-induced immune responses. Furthermore, 54% of COVID-unvaccinated participants in Kilifi and 67% in Nairobi were positive for anti-N IgG, supporting the inference of infection-driven anti-S IgG seroprevalence in this group. Anti-S IgG seroprevalence among COVID-vaccinated individuals appeared to be driven by both infection and vaccination as anti-N IgG seroprevalence in that group was 58% in Kilifi and 79% in Nairobi.

Our findings also indicate temporal increases in SARS-CoV-2 seroprevalence within the general population in Kenya. Anti-S IgG seroprevalence increased more than 3-fold within the Kilifi HDSS site and more than 2-fold within the Nairobi Urban HDSS site since May 2021, when it was 20% and 40%, respectively.^6^ This temporal increase was driven by COVID-19 vaccination rollout beginning in March 2021, as well as by the Delta variant and Omicron BA.1 waves which peaked in August 2021 and December 2021, respectively.^14^ The current seroprevalence estimates from our study are slightly lower than recent estimates from South Africa, a setting with comparable COVID-19 vaccine uptake. Seroprevalence in Gauteng, a predominantly urban province in South Africa, was 73% by December 2021 and 91% by June 2022.^15,16^ It was 95% nationally by March 2022 as estimated using nationally-representative samples from South African blood banks.^17^ Nonetheless, taken together with recent evidence such as that from South Africa, the findings from this study point to high seroprevalence of anti-SARS-CoV-2 IgG in similar settings.

Anti-S IgG seroprevalence and concentrations were significantly lower in Kilifi than in Nairobi. Of note, approximately 25-30% of COVID-unvaccinated adults aged ≥65 years in Kilifi were seronegative for anti-S IgG. These findings suggest a population immunity gap in Kilifi, though this is probably indicative of rural Kenya in general. Anti-S IgG seroprevalence and concentrations were also significantly lower among children aged <16 years than among older age groups. In addition, we found that recent COVID-like symptoms were more likely to be reported among children aged <16 years than in other age groups. Although the burden of severe COVID-19 appears to be lower in children than among adults, as children age into adulthood there may accumulate a significant population immunity gap. Furthermore, a sizeable propoprtion of potentially susceptible children (unvaccinated and with no evidence of an infection-induced SARS-CoV-2 immune response) may substantially contribute to continued transmission within the community.

The relevance of our findings for public health planning was strengthened by use of the WHO IS for anti-SARS-CoV-2 immunoglobulin, which provided an opportunity to estimate anti-S IgG concentrations. We demonstrated that 63% of study participants within the Kilifi HDSS and 86% within the Nairobi Urban HDSS had anti-S IgG concentrations associated with 80% protection against wildtype SARS-CoV-2 among individuals vaccinated using 1-2 doses of mRNA or vectored COVID-19 vaccines. Although identification of correlates of protection for SARS-CoV-2 is intended to inform vaccine development/ licensure, proposed thresholds may inform inferences about population immunity in settings which are characterized by high seroprevalence but low COVID-19 vaccination uptake, such as Kenya. The proportions of study participants with putative protective antibody levels from our analysis may overestimate population immunity if correlates of protection are different for vaccine-induced *vs* infection-induced immune responses or if protective thresholds are higher for future variants of concern. For example, a threshold of 168 BAU/mL has been associated with 80% protection against the alpha variant and thresholds as high as 490 BAU/mL have been proposed for the Delta variant.^13^ Still, as more evidence emerges on correlates of protection, and potentially thresholds relevant for infection-induced immunity, the RCDCs from our analysis may come in useful for estimating population-level protection assuming various antibody concentration cut-offs. We also found that anti-S IgG concentrations were higher among COVID-vaccinated individuals compared to COVID-unvaccinated individuals, underscoring the utility of vaccination for boosting protective antibody levels should higher thresholds of protection be required.

Despite adjustment for test performance, seroprevalence among COVID-unvaccinated individuals as measured using anti-N IgG was substantially lower than anti-S IgG seroprevalence. Anti-N IgG has previously been shown to wane beginning two months post-SARS-CoV-2 infection with sero-reversion within 6 months among about 60% of individuals with mild COVID-19.^18^ The findings from this study indicate that anti-N IgG seroprevalence may underestimate infection-induced immune responses, similar to findings by others.^16,19,20^ Nevertheless, anti-N IgG seroprevalence provided supporting evidence of widespread SARS-CoV-2 infection within the general population in Kenya.

The findings may be subject to some limitations. About 20-30% of the initially randomly sampled individuals were not contactable and were therefore replaced. This may underestimate seroprevalence if individuals typically not found at home were more likely to have been infected with SARS-CoV-2 or to have been vaccinated. Full COVID-19 vaccination coverage was lower in the study sample compared to the respective county-specific estimates from the national COVID-19 vaccination program, therefore the study sample may not be representative of the general population within the respective counties. If COVID-unvaccinated individuals were more likely to have been previously infected, we may have overestimated cumulative incidence. However, we found that anti-N IgG seroprevalence was comparable among COVID-vaccinated and COVID-unvaccinated individuals. The majority of the COVID-19 vaccination data were collected using verbal report. However, as mentioned previously, COVID-vaccinated individuals made up a minority of the study sample, and, in an ongoing exercise, a majority of verbal COVID-19 vaccination reports have been verified against documented vaccination. We did not have anti-N IgG seroprevalence data for 53% of the Nairobi Urban HDSS sample due to delays in testing, which may have introduced bias of unknown direction in anti-N seroprevalence estimates. Finally, anti-S seroprevalence levels may have been underestimated in the primary analyses as they were not adjusted for test-performance.

Based on our findings, we have two key recommendations for policymakers. First, there is a need to support ongoing SARS-CoV-2 serosurveillance to ensure availability of up-to-date seroprevalence data for public health planning. Incorporation of the WHO IS for anti-SARS-CoV-2 immunoglobulin in serosurveillance will be important for informing population-level protection. Second, given a potential population immunity gap in rural Kenya, there is a need to implement interventions to improve uptake of COVID-19 vaccination among rural dwellers at high risk of severe disease, such as the elderly and immunocompromised.

## Data Availability

All data produced in the present study are available upon reasonable request to the authors.

## Acknowledgments

We thank residents of the Kilifi HDSS and the Nairobi Urban HDSS, field staff teams, administrative staff, laboratory staff and community engagement teams who worked on the study. We thank the members of the Kilifi County Health Management Team (CHMT), members of the Nairobi CHMT, participating health facilities and members of the Kenya Multisite Integrated Serosurveillance Steering Committee. This paper is published with the permission of the Director, Kenya Medical Research Institute.

## Disclaimer

The findings and conclusions in this study are those of the authors and do not necessarily represent the official position of the U.S. Centers for Disease Control and Prevention.

## Author contributions

Study conceptualization and design: EWK, IMOA, AKZ, RA, PA, MM, KK, WN, SV, SU, KEG, AOE, GMW, JAGS, AA

Data acquisition: DA, MN, AS, BK, AN

Laboratory measurements and analysis: JN, BK, HK, JG, AK, MT

Data analysis: EWK, BK, AN, MO, CB

Data interpretation: EWK, AKZ, JN, MK, DA, MN, AS, AK, BK, MO, AN, RA, PA, MM, KK, WN, SV, LIO-O, CB, AN, PKM, GB, EM, SU, KEG, AOE, EB, JM, PB, IMOA, GMW, JAGS, AA

Draft preparation: EWK, AKZ, GMW, JAGS, AA

Review and editing: EWK, AKZ, JN, BK, MK, DA, MN, AS, DM, HK, JG, AK, MT, BK, MO, AN, RA, PA, MM, KK, WN, SV, LIO-O, CB, AN, PKM, GB, EM, SU, KEG, AOE, EB, JM, PB, IMOA, GMW, JAGS, AA

