## Supplemental materials for "SARS-CoV-2 seroprevalence and implications for population immunity: Evidence from two Health and Demographic Surveillance System sites in Kenya, February-June 2022"

**Table S1.** Participant characteristics by site

| | Kilifi HDSS | | | Nairobi Urban HDSS | | |
|---|---|---|---|---|---|---|
| **Characteristic** | **N** | **n** | **%** | **N** | **n** | **%** |
| **Sex** | | | | | | |
| Female | 852 | 468 | 54.9 | 851 | 404 | 47.5 |
| Male | 852 | 384 | 45.1 | 851 | 447 | 52.5 |
| **Age category** | | | | | | |
| <16 years | 852 | 313 | 36.7 | 851 | 306 | 36.0 |
| 16-24 years | 852 | 86 | 10.1 | 851 | 94 | 11.0 |
| 25-34 years | 852 | 100 | 11.7 | 851 | 100 | 11.8 |
| 35-44 years | 852 | 100 | 11.7 | 851 | 99 | 11.6 |
| 45-54 years | 852 | 102 | 12.0 | 851 | 101 | 11.9 |
| 55-64 years | 852 | 101 | 11.9 | 851 | 100 | 11.8 |
| >=65 years | 852 | 50 | 5.9 | 851 | 51 | 6.0 |
| **COVID vaccination coverage*** | | | | | | |
| >=1 doses, all ages | 852 | 94 | 11.0 (9.0 – 13.3) | 851 | 284 | 33.4 (30.2 – 36.6) |
| >=1 doses, >=15 years | 559 | 94 | 16.8 (13.8 – 20.2) | 563 | 284 | 50.4 (46.2 – 54.6) |
| Full vaccination, >=15 years | 559 | 55 | 9.8 (7.5 – 12.6) | 563 | 211 | 37.5 (33.5 – 41.6) |
| **History of COVID-like symptoms** | | | | | | |
| >=1 symptoms in the 2 weeks prior to sample collection | 852 | 177 | 20.8 | 851 | 414 | 48.6 |
| Symptoms resulting in hospitalization | 177 | 3 | 1.7 | 414 | 6 | 1.4 |
| **Ever confirmed COVID** | 852 | 4 | 0.5 | 851 | 18 | 2.1 |

*95% confidence intervals indicated in parenthesis

**Table S2.** Proportion of individuals with anti-S IgG concentrations ≥154 BAU/mL by site, sex, age category and COVID vaccination status, and p-values for stratum-specific tests of equality of anti-S concentration distributions.

| KILIFI HDSS | Sample size | n with anti-S IgG ≥154 BAU/mL | Crude proportion with anti-S IgG ≥154 BAU/mL | p-value (equality of distributions test) |
|---|---|---|---|---|
| **Overall** | 852 | 534 | 62.6 | Ref |
| **Sex** | | | | |
| Female | 468 | 232 | 60.4 | Ref |
| Male | 384 | 301 | 64.3 | 0.104 |
| **Age category** | | | | |
| <16 years | 313 | 169 | 54.0 | Ref |
| 16-24 years | 86 | 58 | 67.4 | 0.002 |
| 25-34 years | 100 | 67 | 67.0 | <0.001 |
| 35-44 years | 100 | 67 | 67.0 | <0.001 |
| 45-54 years | 102 | 71 | 69.6 | 0.007 |
| 55-64 years | 101 | 66 | 65.3 | <0.001 |
| ≥65 years | 50 | 35 | 70.0 | 0.063 |
| **COVID vaccination** | | | | |
| Unvaccinated, ≥15 years | 465 | 293 | 63.0 | Ref |
| Vaccinated, ≥15 years | 94 | 88 | 93.6 | <0.001 |
| **NAIROBI URBAN HDSS** | **Sample size** | **n with anti-S IgG ≥154 BAU/mL** | **Crude proportion with anti-S IgG ≥154 BAU/mL** | **p-value (equality of distributions test)** |

| | | | | |
|---|---|---|---|---|
| **Overall** | 851 | 731 | 85.8 | <0.001* |
| **Sex** | | | | |
| Female | 404 | 373 | 83.4 | Ref |
| Male | 447 | 357 | 88.4 | 0.013 |
| **Age category** | | | | |
| <16 years | 306 | 236 | 77.1 | Ref |
| 16-24 years | 94 | 84 | 89.4 | <0.001 |
| 25-34 years | 100 | 84 | 84.0 | <0.001 |
| 35-44 years | 99 | 93 | 93.9 | <0.001 |
| 45-54 years | 101 | 95 | 94.1 | <0.001 |
| 55-64 years | 100 | 93 | 93.0 | <0.001 |
| ≥65 years | 51 | 45 | 88.2 | <0.001 |
| **COVID vaccination** | | | | |
| Unvaccinated, ≥15 years | 279 | 238 | 85.3 | Ref |
| Vaccinated, ≥15 years | 284 | 272 | 95.8 | <0.001 |

*p-value for test of equality of distribution of anti-S IgG concentration cumulative distribution functions at the Kilifi HDSS (ref) *vs* the Nairobi Urban HDSS, overall.

**Table S3.** Test-adjusted **anti-S IgG** seroprevalence (last column) among **COVID-unvaccinated** participants.

| KILIFI HDSS | Sample size | Seropositive | Bayesian population-weighted seroprevalence (95% CrI) | Bayesian population-weighted, test-adjusted seroprevalence (95% CrI) |
|---|---|---|---|---|
| Overall | 758 | 508 | 66.7 (63.3-70.0) | 72.4 (67.6-77.8) |
| **Sex** | | | | |
| Female | 422 | 293 | 69.3 (64.7-73.7) | 75.2 (69.3-81.4) |
| Male | 336 | 215 | 63.8 (58.6-68.7) | 69.3 (62.8-76.0) |
| **Age category** | | | | |
| <16 years | 312 | 194 | 64.4 (59.1-69.0) | 69.9 (63.3-76.4) |
| 16-24 years | 74 | 53 | 68.8 (62.5-76.0) | 74.7 (67.2-83.5) |
| 25-34 years | 86 | 59 | 68.1 (61.6-75.0) | 74.0 (66.5-82.0) |
| 35-44 years | 88 | 63 | 69.2 (63.1-76.1) | 75.1 (67.7-83.6) |
| 45-54 years | 81 | 58 | 69.1 (62.8-76.3) | 74.8 (67.4-83.3) |
| 55-64 years | 75 | 51 | 68.0 (61.1-74.6) | 73.8 (65.8-81.9) |
| ≥65 years | 42 | 30 | 68.9 (61.7-77.0) | 74.6 (66.3-84.0) |

| NAIROBI URBAN HDSS | Sample size | Seropositive | Bayesian population-weighted seroprevalence (95% CrI) | Bayesian population-weighted, test-adjusted seroprevalence (95% CrI) |
|---|---|---|---|---|
| Overall | 567 | 479 | 85.3 (82.1-88.2) | 93.8 (88.2-99.7) |
| **Sex** | | | | |
| Female | 283 | 248 | 88.5 (84.5-92.0) | 97.4 (91.1-100) |
| Male | 284 | 231 | 82.5 (77.8-86.7) | 90.6 (84.0-99.9) |
| **Age category** | | | | |
| <16 years | 300 | 242 | 81.4 (76.8-85.5) | 91.7 (83.7-99.7) |
| 16-24 years | 60 | 52 | 86.8 (79.7-92.7) | 95.4 (88.2-99.8) |
| 25-34 years | 65 | 55 | 85.8 (78.7-91.8) | 94.0 (86.2-99.8) |
| 35-44 years | 48 | 45 | 89.7 (82.5-95.9) | 95.6 (89.2-99.8) |
| 45-54 years | 40 | 35 | 86.3 (77.7-93.3) | 93.9 (85.3-99.8) |
| 55-64 years | 39 | 38 | 90.5 (82.8-97.1) | 95.4 (88.8-99.8) |
| ≥65 years | 15 | 12 | 84.7 (72.0-93.2) | 94.2 (84.3-99.8) |

**Figure S1.** Population density map showing the geographical location of study sites within Kenya. Dots represent 100 population.

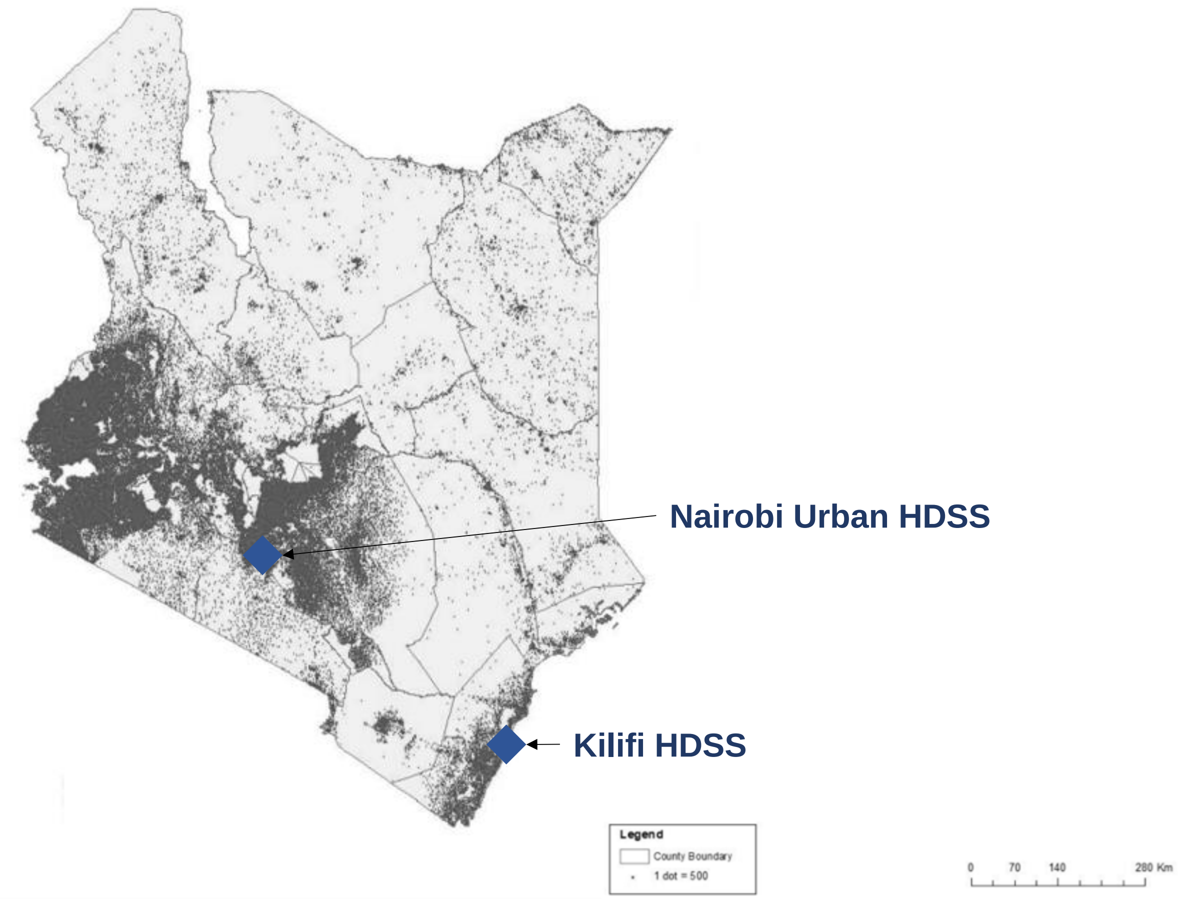

**Figure S2.** Site-specific enrolment periods overlaid on daily number of confirmed cases

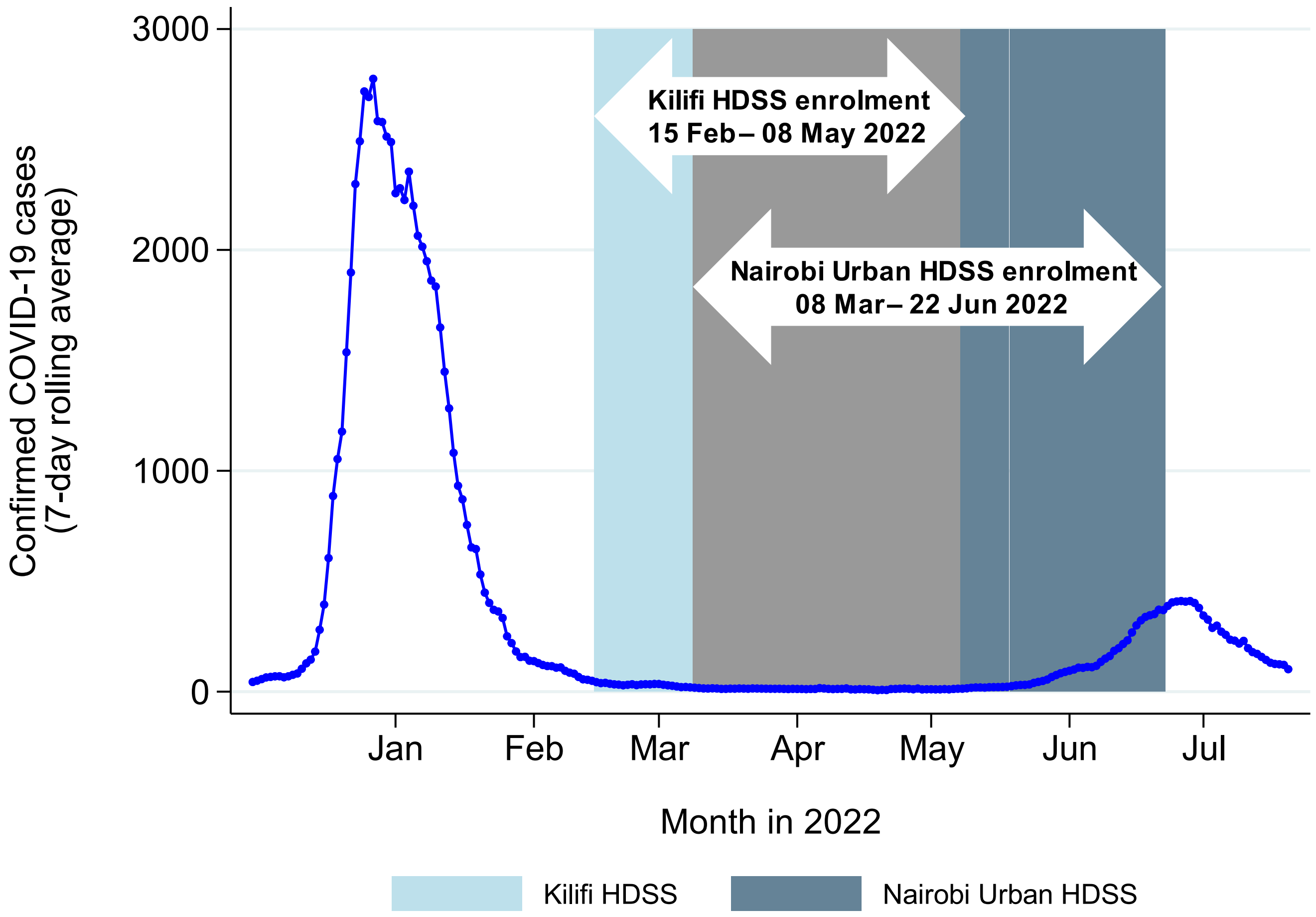

**Figure S3.** Frequency of COVID-like symptoms within the two weeks prior to sample collection overall.

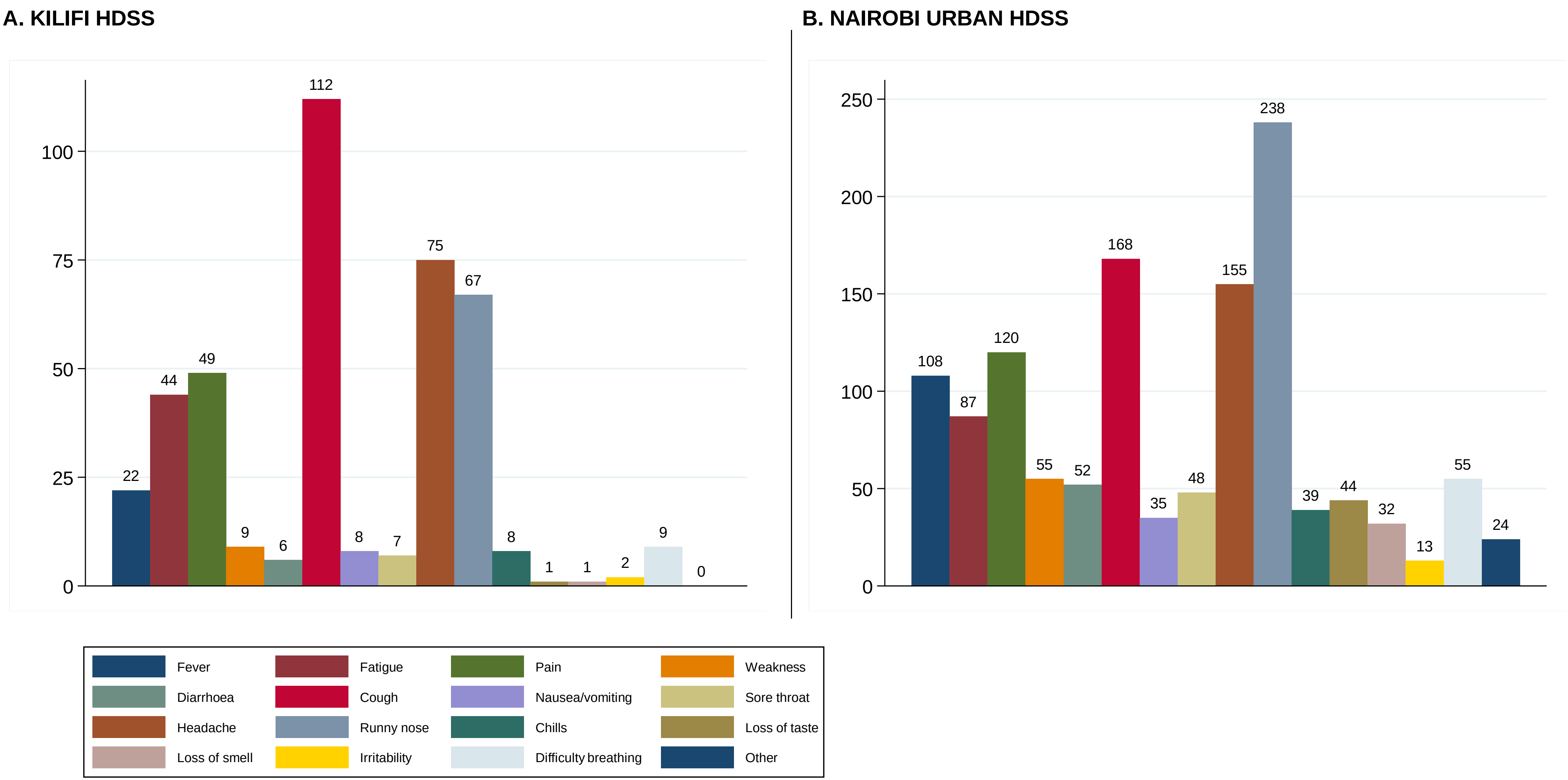

**Figure S4.** Frequency of COVID-like symptoms within the two weeks prior to sample collection by age category.

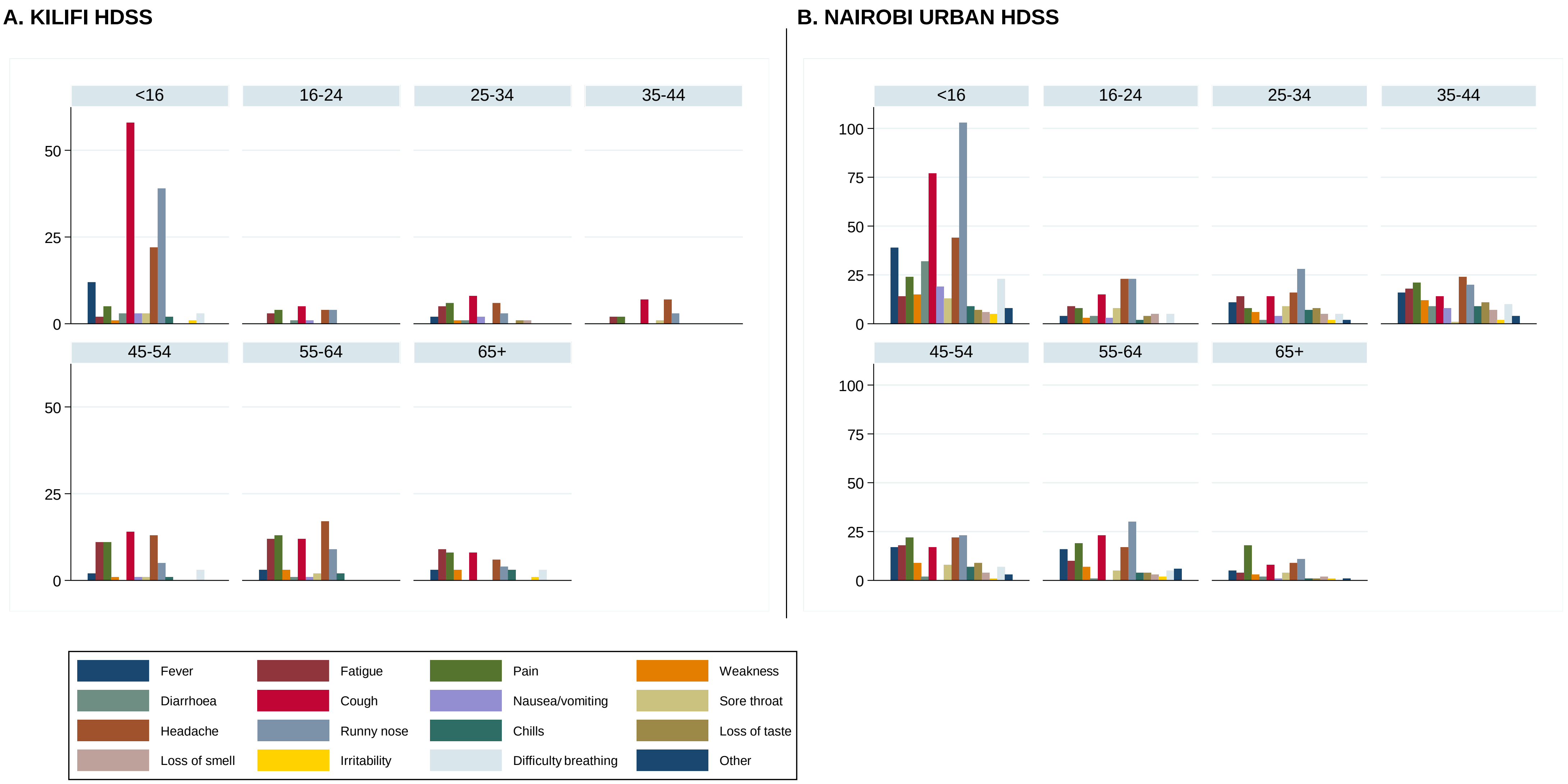

**Figure S5.** Reverse cumulative distribution curves of anti-S IgG concentrations within the Kilifi HDSS and Nairobi Urban HDSS by sex (A,D), by age category (B, E) and by COVID vaccination status among individuals aged ≥15 years (C, F). The red vertical line represents an antibody concentration of 154 BAU/mL.

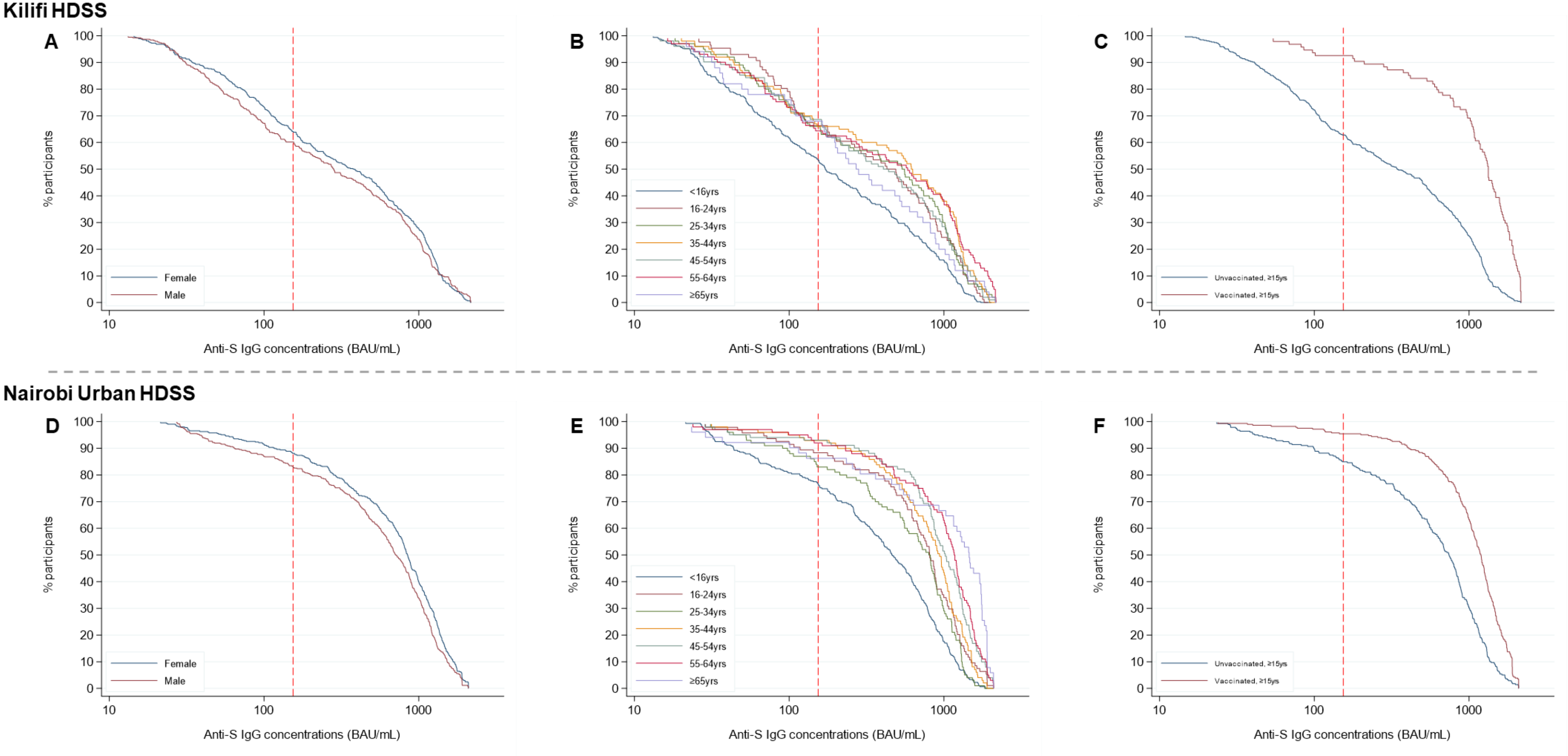

### Study data collection form

| | | |
|---|---|---|
| **Study:** S-COV-2 HDSS sero-surveys | **1. Location (LOC):**<br>□**KILIFI- KLF**<br>□**NAIROBI-NRB** | **2.1 Participant PID:**_____________<br>**2.2 Participant ID: (SS-\| \| \| - \|___\|___\|___\|___\|**<br>**LOC S/N** |
| **3. Date of visit (DD/MMM/YYYY): \|___\|___\| / \|___\|___\|___\| / \|___\|___\|___\|___\|** | | **4. Staff initials: \|__\|__\|__\|** |

| | | |
|---|---|---|
| **5.** | **5.1** Residency status:<br>a. Present<br>b. Not at Home<br>c. Await Husband Decision<br>d. Guardian Not Present<br>e. Out Migrated<br>f. Died<br>g. Not Known | **5.2** Consent: \|__\| (0= No , 1=Yes, 2=Withdrawn)<br>**5.3** If Consent is NO/Withdrawn: Reason why refused/Withdrawn to participate .<br>a. Parent/Guardian refused consent for the study<br>b. Mother needs the father to give consent and the father is away<br>c. Fear of blood/Swab<br>d. Not interested in the research<br>e. Negative attitude towards KEMRI/ APHRC and KEMRI / APHRC activities<br>f. Prior experience with KEMRI/ APHRC<br>g. Others |
| **6.** | **6.1** First name: ___________________________________<br>**6.2** Middle name: ________________________________<br>**6.3** Surname:_____________________________________ | **6.4** Education: \|__\| (Fill 0=No formal education; 1=Incomplete primary; 2= Complete primary, 3=Incomplete secondary; 4=Complete secondary; 5= Post secondary)<br>**6.5** Religion: \|__\| (0=None, 1= Muslim 2= Catholic, 3=Protestant; 4= Other Christians, 5= Traditional religion, 6=Hindu, 7=Other(specify).................................) |
| **7.** | **7.1** Estimated age in years: \|__\|__\|__\| | **7.2** if <5 years, DOB \|__\|__\|/\|__\|__\|__\|/\|__\|__\|<br>(dd/MMM/yy) |
| **8.** | Sex at birth: | **\|**__\| (Fill in 0= Female , 1=Male, 2=Other) |
| **9.** | Usual place of residence (**NOT ancestral home**) | |
| | **9.1** County: _______________________<br>**9.2** Sub-county: ____________________<br>**9.3** Division: _______________________ | **9.4** Location: _______________________<br>**9.5** Sub-location: ___________________<br>**9.6** Village/estate: __________________ |

<table>
<tr><td>**10.0**</td><td colspan="2">**10.1 In the past 3 months, have you experienced any of the following** (check all reported symptoms):<br>□ History of fever :**|__| 0= No ; 1=Yes**<br>□ Chills :**|__| 0= No ; 1=Yes** □ Shortness of breath/difficulty breathing □ Pain (check all that apply)<br>□ General weakness □ Diarrhoea:**|__| 0= No ; 1=Yes** *( ) Muscular ( ) Chest*<br>□ Cough:**|__| 0= No ; 1=Yes** □ Nausea/vomiting:**|__| 0= No ; 1=Yes** *( ) Abdominal ( ) Joint*<br>□ Sore throat:**|__| 0= No ; 1=Yes** □ Headache:**|__| 0= No ; 1=Yes**<br>□ Congestion/Runny nose:**|__| 0= No ; 1=Yes** □ Irritability/Confusion :**|__| 0= No ; 1=Yes**<br>□ Fatigue:**|__| 0= No ; 1=Yes-** □ UNKNOWN (all)<br>□ New loss of taste :**|__| 0= No ; 1=Yes** □ Other, specify<br>□ New loss of smell :**|__| 0= No ; 1=Yes** □ NO SYMPTOMS<br>□ Pain (check all that apply): ( )Muscular; ( )Chest; ( )Abdominal ( )Joint</td></tr>
<tr><td></td><td colspan="2">**10.1** Which of the above symptoms(**Q10.1**) have you experienced in the **last 2 weeks:{ } Choose all that apply from Q10**</td></tr>
<tr><td>**11.**</td><td>If you answered yes, Q10,</td><td>**11.1** Did any of these symptoms require you to seek medical attention?<br>**|**__| (Fill in 0= No , 1=Yes, 2= Don't know)<br><br>**11.2** Did any of these symptoms cause you to miss work?<br>**|**__| (Fill in 0= No , 1=Yes, 2= Don't know)<br><br>**11.3** Did any of these symptoms require you to be hospitalised?<br>**|**__| (Fill in 0= No , 1=Yes, 2= Don't know)</td></tr>
<tr><td>**12.**</td><td>COVID-19 vaccination status</td><td>12.1 Have you had a COVID-19 vaccine? |__| (Fill in 0= No , 1=Yes, 2= Don't know)<br><br>COVID vaccine type? (1 – AstraZeneca, 2 – Pfizer, 3- Johnson & Johnson, 4 -Moderna, 5 - SputnikV, 6-Sinopharm ; 7- Others………….(Type), 8=Don't Know)<br><br>12.2 How many doses? |__| (1 dose=1, 2 doses =2, 3 doses-3)<br>a) **Dose 1 type:** |__|date |_d_|_d_|/|_m_|_m_|_m_|/|_y_|_y_|<br>**b) Dose 2 type:** |__| date |_d_|_d_|/|_m_|_m_|_m_|/|_y_|_y_|<br>**c) Dose 3 type:** |__|date |_d_|_d_|/|_m_|_m_|_m_|/|_y_|_y_|<br>**12.3 Vaccination confirmed by: | | 1=Vaccination Card; 2= SMS; 3= Verbal report; 4=Other(specify)..........................**</td></tr>
<tr><td>**13.**</td><td>Check for BCG scar (first look at the left, then the right shoulder)</td><td>**13.1** Does the participant have a BCG scar? **|**__| (Fill in 0= No , 1=Yes, 2= Don't know)</td></tr>
<tr><td>**14.**</td><td>For children < 5 years, vaccination information from health card</td><td>**14.1** Do you have vaccination records for *(child's Name)*? **|**__| (Fill in 0= No , 1=Yes)<br>If you answered YES, in 20.1, *Capture Vaccination history from the records provided* (Fill in 0= No , 1=Yes, 2= Don't know)<br>**14.2** BCG **|**__| if yes, fill in date |_d_|_d_|/|_m_|_m_|_m_|/|_y_|_y_|<br>**14.3** Penta 1 **|**__| if yes, fill in date |_d_|_d_|/|_m_|_m_|_m_|/|_y_|_y_|<br>**14.4** Penta 2 **|**__| if yes, fill in date |_d_|_d_|/|_m_|_m_|_m_|/|_y_|_y_|<br>**14.5** Penta 3 **|**__| if yes, fill in date |_d_|_d_|/|_m_|_m_|_m_|/|_y_|_y_|<br>**14.6** PCV 1 **|**__| if yes, fill in date |_d_|_d_|/|_m_|_m_|_m_|/|_y_|_y_|<br>**14.7** PCV 2 **|**__| if yes, fill in date |_d_|_d_|/|_m_|_m_|_m_|/|_y_|_y_|<br>**14.8** PCV 3 **|**__| if yes, fill in date |_d_|_d_|/|_m_|_m_|_m_|/|_y_|_y_|<br>**14.9** MCV1 **|**__| if yes, fill in date |_d_|_d_|/|_m_|_m_|_m_|/|_y_|_y_|<br>**14.10** MCV2 **|**__| if yes, fill in date |_d_|_d_|/|_m_|_m_|_m_|/|_y_|_y_|<br>If you answered NO, in 20.1,<br>**14.11** Reason why vaccination card is not available. **|**__|<br>*(Choose from the List of probable reasons why no records provided )*<br>1. Mother/guardian who keeps the card is not available<br>2. Claims to have had a card but misplaced it<br>3. Was not issued with the card at the health facility but received vaccinations<br>4. Has never gotten a card and never received vaccinations<br>5. Card Destroyed/burnt<br>6. Card left at another place other than the current residence</td></tr>
<tr><td>**15**</td><td>Have you ever tested for COVID?*)*?<br>**|**__| (Fill in 0= No , 1=Yes)</td><td>If Positive , Date of last positive test |d_|d_|_m_|_m_|/|_y_|_y_|</td></tr>
</table>

| | | |
|---|---|---|
| **16.** | Time of sample collection (hh:mm; 24hr clock) | _____:____ or \|__\| N/A sample not taken |

**Form completed by**: ____ ____________________ _(Signature) |__|__|/|__|__|__|/|_2_|_0_|__|__ |(Date (dd/MMM/yyyy))
